# Maternal PrEP use in HIV-uninfected pregnant women in South Africa: Role of Stigma in PrEP initiation, retention and adherence

**DOI:** 10.1101/2020.11.24.20237867

**Authors:** Alexander Moran, Nyiko Mashele, Rufaro Mvududu, Pamina Gorbach, Linda-Gail Bekker, Thomas J. Coates, Landon Myer, Dvora Joseph Davey

## Abstract

Pregnant women in sub-Saharan Africa are at high risk of HIV acquisition and require effective methods to prevent HIV. In a cohort of pregnant women offered pre-exposure prophylaxis (PrEP), we evaluate the relationship between internalized and anticipated stigma and PrEP initiation at first antenatal visit, 3-month continuation and adherence using multivariable logistic regression. High internalized and anticipated PrEP stigma are associated with lower PrEP initiation at first antenatal visit (aOR internalized stigma=0.06; 95%CI=0.03-0.12 & aOR anticipated stigma=0.53; 95%CI=0.29-0.97) compared to women with low reported stigma, after controlling for covariates. Women whose partners have not been tested for HIV or whose serostatus remains unknown have 1.6-times odds of PrEP retention at 3-months compared to women whose partners have been tested (aOR=1.60; 95%CI=1.02-2.52) after adjusting for covariates. PrEP counseling and maternal PrEP interventions must consider individual- and relational-level interventions to overcome anticipated PrEP stigma and other barriers to PrEP initiation and adherence.

## Introduction

HIV incidence among pregnant women in South Africa remains high despite improved initiation of antiretroviral therapy (ART), and seroconversion during pregnancy or the post-partum period continues to contribute to pediatric HIV incidence. (1–3) Aside from hormonal changes which may affect HIV risk, behavioral changes during pregnancy and the post-partum period remain important determinants of HIV acquisition, including condom use, condom negotiation, partner HIV testing and sexual activity. (1,4–7) These risks are compounded among pregnant adolescent girls and young women in South Africa, who may have lower HIV care engagement and later, or less frequent, antenatal care (ANC) attendance. (8) HIV pre-exposure prophylaxis (PrEP) is a safe, effective and woman-controlled HIV prevention method. (9–11) PrEP is effective in preventing HIV transmission in many settings including heterosexual HIV transmission and transmission among men who have sex with men (MSM). (12–15) Despite these successes, continuation on and adherence to PrEP remain challenges, especially among pregnant and postpartum women. (16,17)

There is a dearth of research on stigma and its association with PrEP initiation, continuation and adherence among pregnant women in South Africa. Both anticipated and internalized stigma may serve as important barriers to PrEP initiation, continuation on PrEP and good PrEP adherence among pregnant and postpartum women. (18–22) Anticipated stigma, which describes the expectation of prejudicial or discriminatory behavior because of PrEP use, and internalized stigma, which describes an individual’s own belief in the negative ideas associated with PrEP, may act simultaneously or independently. (23,24)

We did this study to understand the role of anticipated and internalized stigma in PrEP initiation, continuation and adherence among pregnant and post-partum women in South Africa. We hypothesize that internalized stigma and anticipated stigma are associated with lower PrEP initiation, continuation and adherence and stand as barriers for pregnant PrEP users. Additionally, we hypothesize that partner HIV serostatus and prior HIV testing are important factors in determining both anticipated stigma and PrEP initiation, continuation and adherence. These findings can inform PrEP delivery strategies, including counseling methods, awareness building in the community, male partner involvement and adherence support strategies.

## Methods

The PrEP-PP (PrEP in Pregnant and Postpartum women) study is an open prospective cohort which enrolls consenting pregnant, HIV-uninfected adolescent girls and women (age >=16 years) at the first antenatal care (ANC) visit and follows participants through 12-months post-delivery (Clinical Trial Registry: NCT03902418). The study recruits at one public health clinic in Cape Town, Western Cape, South Africa. Recruitment began in August 2019 and is ongoing (N=623), with a planned sample size of N=1200 women. The study was approved by the Human Research Ethics Committee at the University of Cape Town (#297/2018) and by the University of California, Los Angeles Institutional Review Board (IRB#18-001622).

### Study Participants

Study eligibility criteria include: 1) age of at least 16 years, 2) confirmed HIV-negative serostatus by a 4^th^ generation antigen/antibody combination HIV test (Abbott Laboratories, Chicago, IL, US) 3) intention of giving birth in the midwife obstetrics unit (MOU) of the enrollment facility, 4) confirmed pregnancy status, 5) absence of psychiatric or medical contraindications to PrEP. Participants are ineligible if any of the following criteria are met: 1) concurrent enrollment in another HIV-1 vaccine or prevention trial, 2) medical hospitalization in the past year for any reason, 3) active TB disease or TB treatment in the past 30 days, 4) history of renal disease, 5) psychotic symptoms or current or past use of anti-psychotic medication, 6) positive Hepatitis B surface antigen (HBsAg) test at initial screening with a rapid test (Abbott Laboratories), 7) history of bone fracture not related to trauma, or 8) any other medical, psychiatric or social condition in which, in the opinion of the investigators, would affect the ability to consent or participate in the study. Participants are censored upon HIV seroconversion, pregnancy loss or infant death, migration away from the study area, transfer out of care at the study facility or loss to follow up (defined by not returning to the study for a clinical visit for more than 90 days after unsuccessful participant tracking by the study staff, or if the participant withdraws consent for future visits).

Health care providers at study facilities provide group counseling at baseline, which includes information on HIV testing and counseling, antiretroviral therapy (ART) for prevention of mother to child transmission of HIV (PMTCT) and the importance of HIV prevention for women who are HIV-negative. Eligible, consenting participants receive 120 Rand (approximately $8 USD) in grocery vouchers for their time and effort in the study, as well as remuneration for transportation costs. Participants also receive refreshments (i.e., sandwiches and a cooldrink) on the day of their visit.

### Data Collection

Following South African HIV testing guidelines, HIV counselors provide all ANC attendees with pre-test counseling for HIV, rapid HIV testing and post-test counseling. (25) Upon confirmation of HIV seronegative status, trained study staff approach women to introduce the HIV prevention study. Upon agreement to participate in the study, the participant consents to screening for study eligibility, which includes a rapid HIV antigen/antibody test and an HBsAg test (Abbott Laboratories). Upon confirmation of eligibility and unassisted study consent, participants complete the baseline visit survey, which takes 30 to 45 minutes using REDCap, a secure, web-based database platform. (26,27) Participants also receive individual counseling about HIV prevention in pregnancy, including PrEP, along with information on consistent and correct condom use, knowing her partner’s HIV status (including referral for male partner or couples HIV testing and counseling), and risk of serodiscordance. At baseline, participants self-collect a vaginal swab that is tested for *Chlamydia trachomatis* (CT), *Neisseria gonorrhoeae* (NG), and *Trichomonas vaginalis* (TV) using point of care testing (Cephid, Inc., Sunnyvale, CA, US) and treatment is received during the same visit following National STI Guidelines. (28) Participants who are diagnosed with a STI receive a partner notification letter for partner STI treatment.

Following the baseline survey, the study interviewer provides information about what PrEP is, and benefits of taking PrEP. The interviewer then asks the participant if they are interested in starting PrEP and if they are unsure or uninterested, this will not impact on their study participation. For study participants who consent to taking PrEP, the study nurse draws blood to measure baseline creatinine levels, results for which are confirmed within 24-48 hours. Upon confirmation that the participant wants to use PrEP, the nurse provides the patient with a one-month supply of Truvada® (tenofovir disoproxil fumarate/emtricitabine [TDF-FTC]) and an invitation card to return in one month for follow up testing (after which participants will receive a three month prescription to correspond with quarterly study follow-up visits). Participants who do not start PrEP receive an invitation to return in three months for a quarterly study follow-up visit. The entire baseline visit lasts between 60 to 90 minutes.

Follow up visits are quarterly and coincide with ANC visits until birth and first postpartum visit. Follow up visits follow Southern African PrEP guidelines and additionally include a follow up visit, adherence counseling for PrEP (if applicable), a blood draw for dried blood spot (DBS) testing to measure serum tenofovir diphosphate (TFV-DP) levels (if PrEP use reported in past 30 days) as a proxy for PrEP adherence (29), and renal function. (30) Follow up visits last approximately 30 to 45 minutes.

### Survey Measures

Survey measures are collected at baseline and follow up visit for all participants (on PrEP and not on PrEP). Survey measures were written in English, translated into isiXhosa, and back translated to English to ensure appropriate translations. Survey measures include questions on: (a) basic demographic information and obstetric history (baseline only), (b) partner HIV status, (c) sexual behaviors in the past month and past week (including number of sex partners, type of sex, frequency of sex and condom use), (d) substance use from the Alcohol Use Disorders Identification Test (AUDIT) (31) and Drug Use Disorders Identification Test (DUDIT) (32), (e) HIV risk perception, (f) intimate partner violence (using the WHO IPV scale (33,34)), (g) perceived partner, community and social support for PrEP (including PrEP and HIV stigma measures), and (h) for PrEP users only, questions related to PrEP adherence according to self-report (seven-day and 30-day recall) and pill count measures, side effects, adverse events, severe adverse events and birth outcomes (after participants have given birth) at follow-up visits.

### PrEP Initiation, Retention, Continuation and Adherence

We defined PrEP initiation as accepting the initial one-month PrEP prescription at baseline. Participants on PrEP at baseline are invited to return for a one-month follow-up visit to monitor adherence and a prescription for a two-month supply of PrEP. PrEP retention was measured at the three-month follow up visit, in which retention was defined by returning for the first quarterly follow up visit, excluding participants who had been censored prior to the three-month follow up visit. We defined PrEP continuation as having returned for the first quarterly follow up visit and having received a PrEP prescription both at the baseline visit and at the one-month follow up visit. Among those who continued on PrEP, PrEP adherence was measured using the 30-day recall self-report measure, in which good adherence was defined by taking at least 25 of the last 30 doses preceding the three-month follow up visit (Figure 1). In sensitivity analyses, we did not find differences between the 7-day and 30-day self-report adherence measure, and in the absence of available DBS TFV-DP results, we report the 30-day self-report measure in favor of the 30-day pill count measure, which had high missingness and which had lower agreement with both self-report measures.

**Figure 1:**
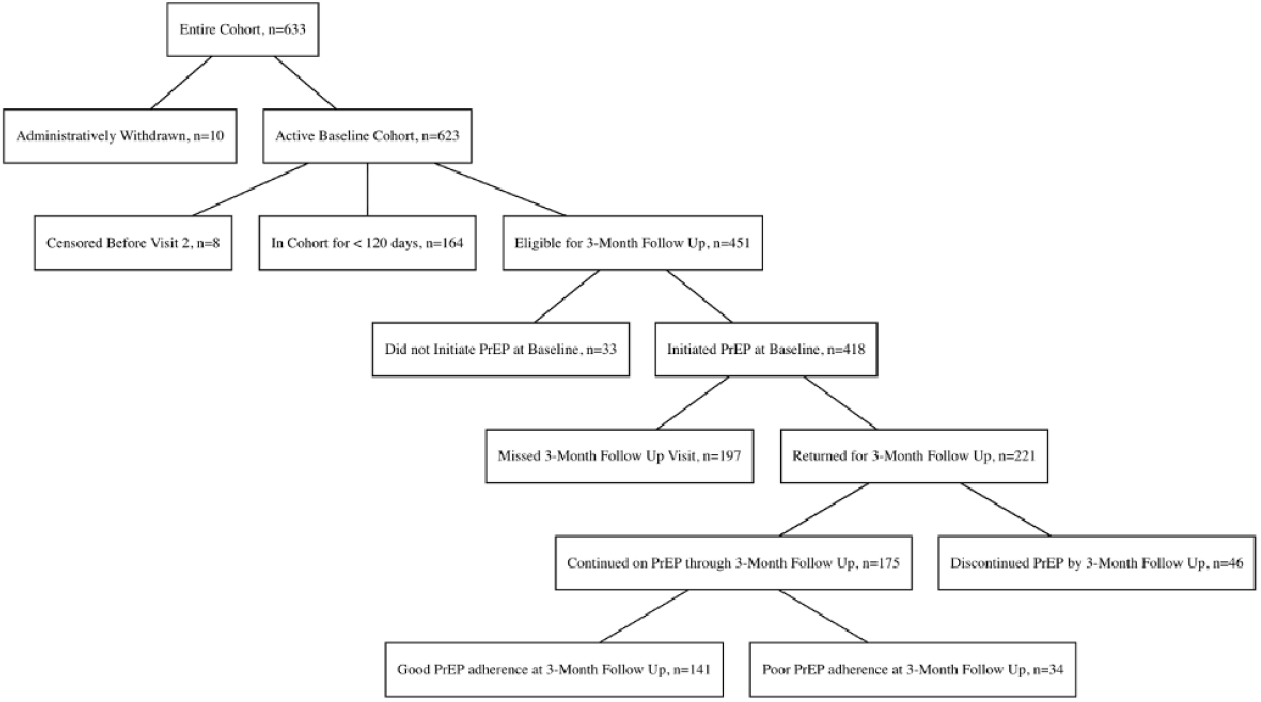
PrEP-PP Cohort as of 9-September 2020.

### PrEP Stigma Scales

We performed an exploratory factor analysis to define two PrEP stigma scales: internalized PrEP stigma and anticipated PrEP stigma. We examined these factors as two separate scales to understand the separate relationships between internalized PrEP stigma and anticipated PrEP stigma, which may function differently in the context of partner HIV testing. Scale measures were dichotomized to high and low stigma, where high stigma indicates that a participant reported a summary stigma score above the median (median =3, range = 0-12 for both internalized PrEP stigma and anticipated PrEP stigma).

### Statistical Analyses

As of 9-September 2020, the study had recruited n=633 participants, of whom 10 were administratively withdrawn because they were ineligible after consenting. We considered the remaining n=623 participants for the PrEP initiation outcome. Among these, n=8 were censored before the three month visit (visit 2), n=164 had been in the cohort for under 120 days without having completed a three-month follow up visit and n=33 did not start PrEP at baseline; these participants were excluded from PrEP retention, continuation and adherence outcomes. Among the n=418 participants on PrEP who had been in the cohort for at least 120 days, we considered all for the PrEP retention outcome. We measured PrEP continuation (receiving a PrEP prescription at both baseline and the one-month follow up visit) among the n=221 participants who attended the first quarterly follow up visit. We finally measured adherence among the n=175 participants who continued on PrEP through the first quarterly follow up visit (Figure 1).

We present distribution of PrEP initiation (PrEP prescription received or not at baseline), PrEP retention at three months (returned to the three-month follow up visit or missed the visit), PrEP continuation (received a PrEP prescription at both baseline and the one-month refill visit) and PrEP adherence at three months among those who continued (good adherence – defined as having taken at least 25 of the past 30 doses of PrEP, measured by self-report – or poor adherence – defined as having taken fewer than 25 of the past 30 doses of PrEP, measured by self-report), including counts and percentages for categorical variables and medians and interquartile ranges (IQRs) for continuous variables. We note the sample size for each characteristic presented to highlight any missingness. We present demographic characteristics including age, education, income, household size including children, number of living children and marital status. We also include gestational age in weeks at baseline, gravidity (number of prior pregnancies), pregnancy intention and feelings on having a baby. We present HIV risk and sexual behavior data, including partner HIV testing behaviors, partner HIV status, HIV risk perception, sex frequency in the past three months and number of sexual partners during pregnancy. We present self-reported stigma characteristics including internalized PrEP stigma, anticipated PrEP stigma, depression (measured through the Edinburgh Postnatal Depression Scale [EPDS]) (35), concern about sexual violence or rape and IPV experience (in general and emotional, physical or sexual IPV). Finally, we present selected substance use characteristics including any drug or alcohol use in the last year before pregnancy using the AUDIT and DUDIT scales.

We assessed potential confounders with directed acyclic graphs (DAGs) (Supplemental Material). We examined crude associations between possible confounders (gestational age at baseline, education level, gravidity, partner HIV testing, STI at baseline), exposures (internalized PrEP stigma and anticipated PrEP stigma) and outcomes of interest (1. PrEP initiation at baseline, 2. PrEP retention at three months, 3. PrEP continuation at three months and 4. PrEP adherence at three months) in separate models using logistic regression. We considered exact logistic regression but did not find significant differences in the results. We then constructed two separate multivariable logistic regression models: one for internalized PrEP stigma (which controlled for gestational age, education level and gravidity) and one for anticipated PrEP stigma (which controlled for gestational age, education level, gravidity and partner HIV testing) and tested these models for the first three outcomes of interest. For the fourth outcome of interest (PrEP adherence at three months) we also controlled for STI at baseline in both the internalized PrEP stigma model and the anticipated PrEP stigma model. All statistical analyses were conducted with R v4.0 (Vienna, Austria). (36)

## Results

### Descriptive Analyses of Study Baseline

We enrolled and followed 623 eligible pregnant women at their first antenatal care visit. Median age in years at baseline was 25 (IQR: 22-30) and median gestational age at baseline was 21 (IQR: 14-29). Half of participants had less than Grade 12 education (n=313, 50%). Overall, 19% (n=121) were married and 35% (n=218) were cohabiting at baseline, 8% (n=50) reported not being in a relationship. Median gravidity was two (IQR: 1-3) and 35% (n=218) were primigravida. Most participants did not intend to get pregnant or changed their intention (n=428, 69%). Over one-quarter of participants reported that their partner had not tested for HIV or they did not know their partner’s serostatus (n=170, 27%) and reported HIV seropositivity among partners was low (n=8, <2% for all women). Nearly all participants had at least one sexual partner during pregnancy (n=605, 97%) and over half of participants in all samples reported 2-4 sexual events per month in the last three months. STI prevalence of CT, NG and/or TV was 35% (n=216) at first antenatal visit. Reported depression (EPDS score >=13) was 7.2% (n=45) at baseline. Half of participants reported no concern about sexual violence or rape in the next three months, but one-third (n=205, 33%) reported that they were “very concerned”. About one tenth of participants (n=75,12%) reported any kind of IPV in the past year. Almost half of participants (n=292, 47%) reported alcohol and/or drug use in the last 12 months (Table I).

**Table 1.**
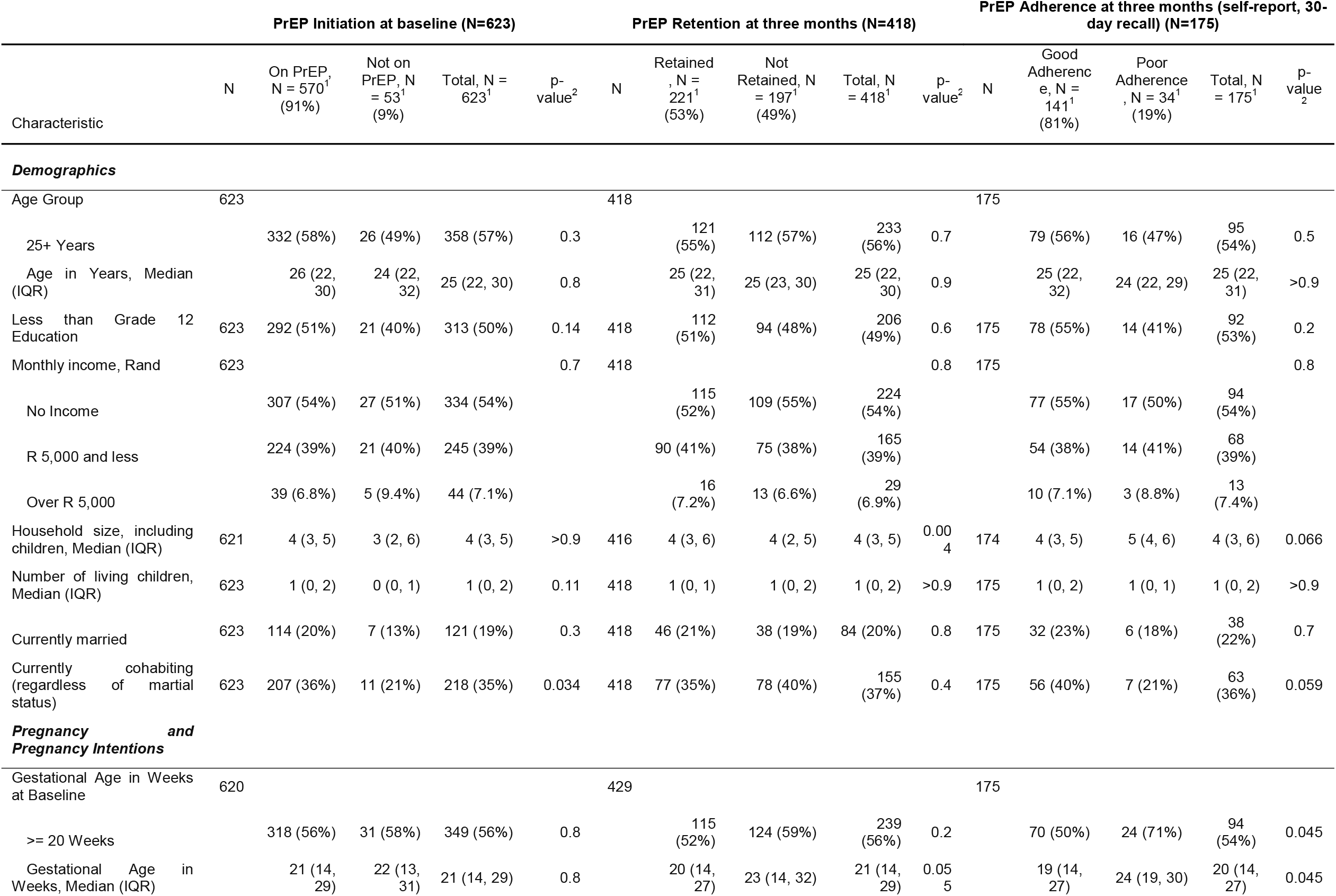
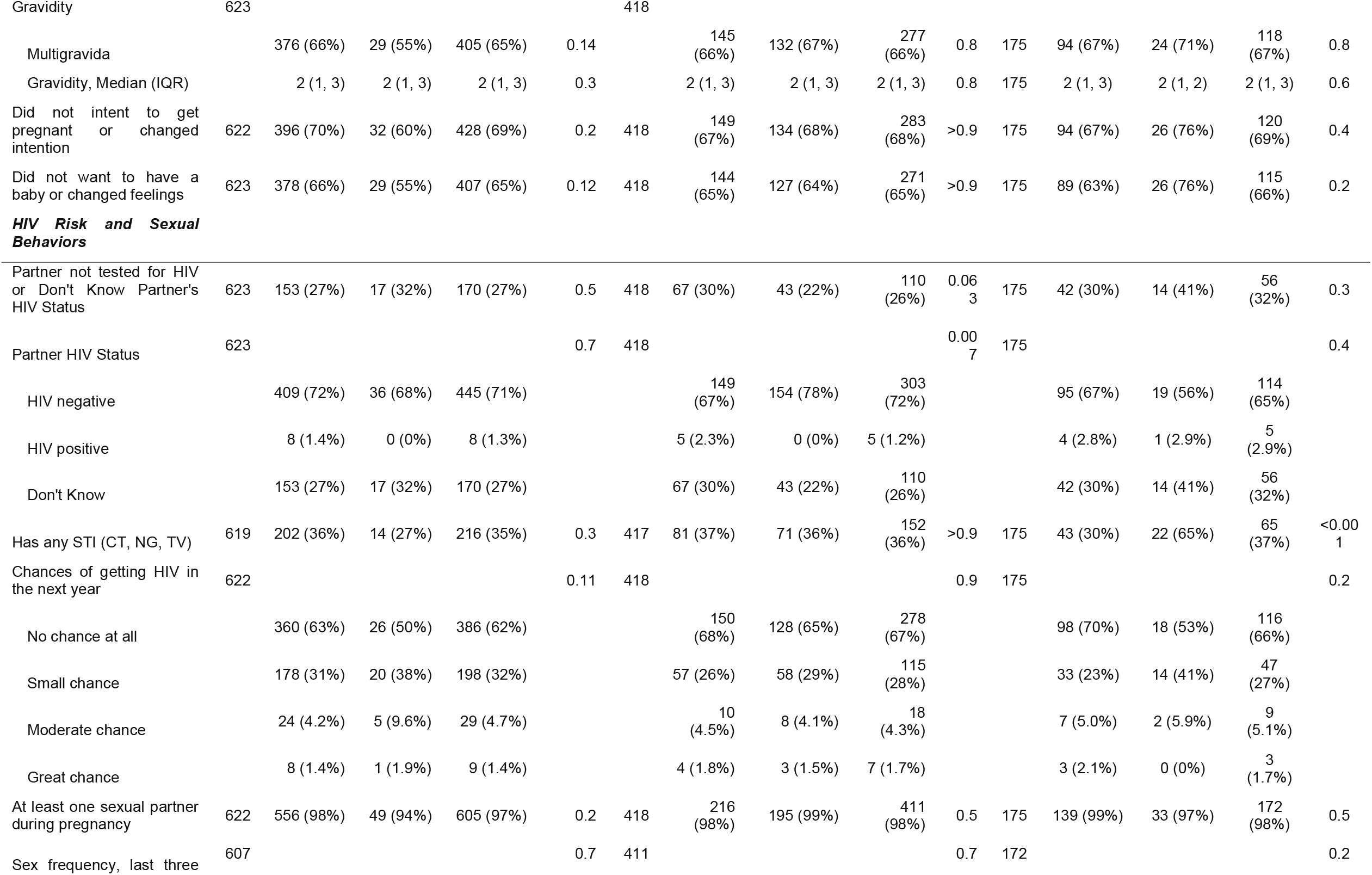
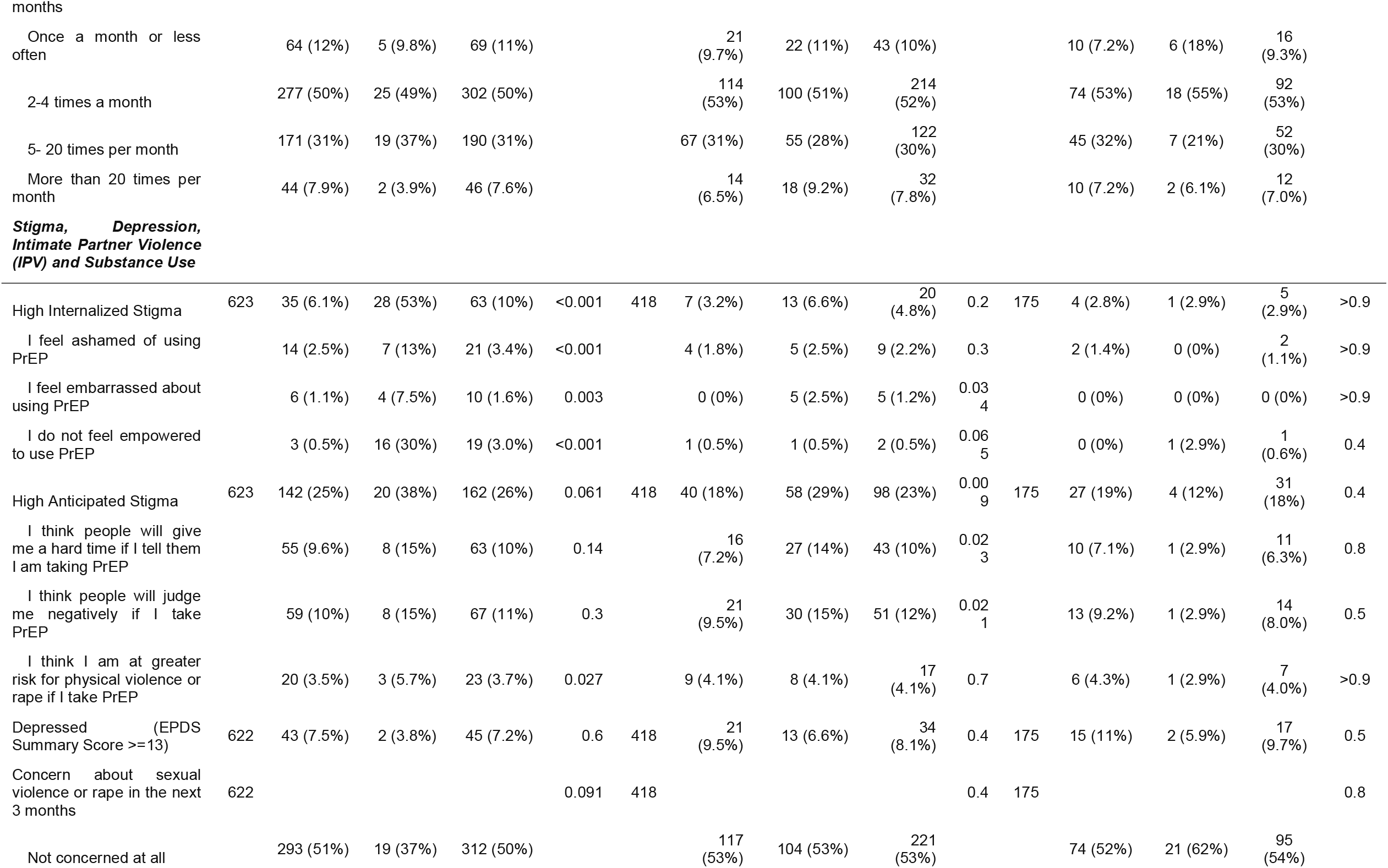
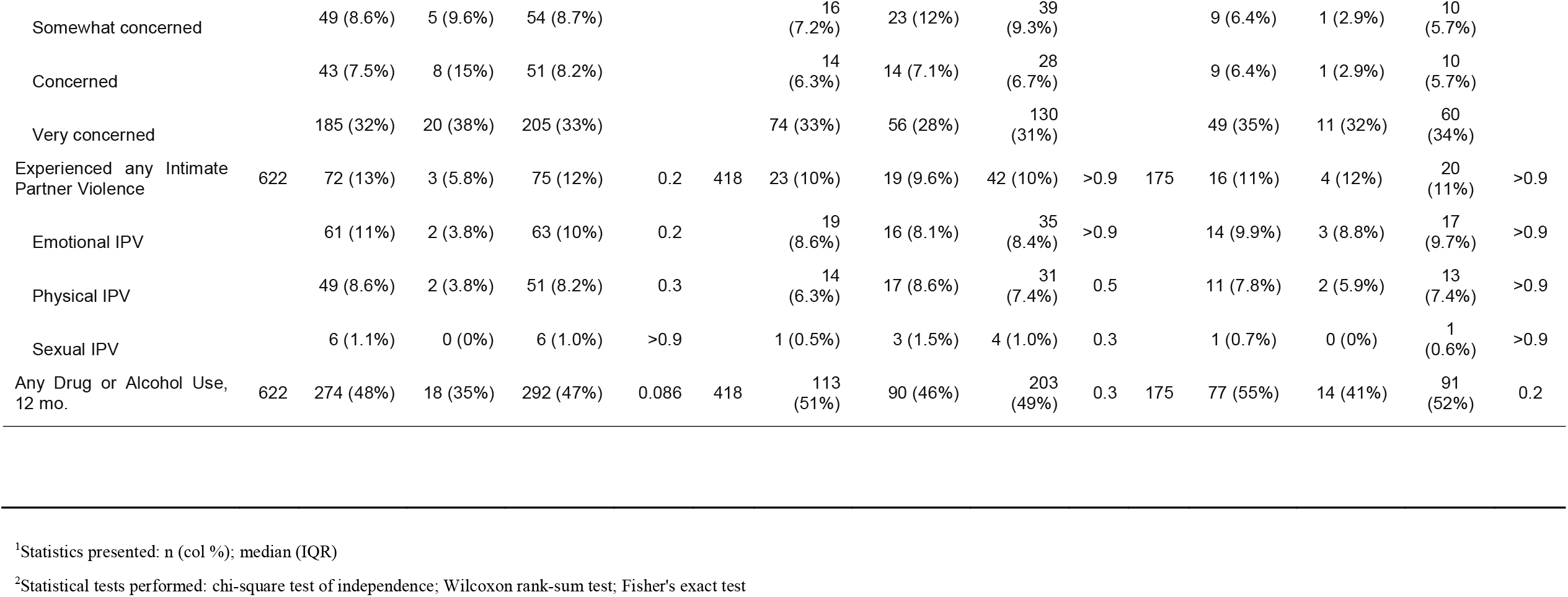
Baseline demographic characteristics of women in the PrEP-PP Study in Cape Town, South Africa, 2019-2020.

| Characteristic | PrEP Initiation at baseline (N=623) |  |  |  |  | PrEP Retention at three months (N=418) |  |  |  |  | PrEP Adherence at three months (self-report, 30-day recall) (N=175) |  |  |  |  |
| --- | --- | --- | --- | --- | --- | --- | --- | --- | --- | --- | --- | --- | --- | --- | --- |
|  | N | On PrEP, N = 570<sup>1</sup> (91%) | Not on PrEP, N = 53<sup>1</sup> (9%) | Total, N = 623<sup>1</sup> | p-value<sup>2</sup> | N | Retained, N = 221<sup>1</sup> (53%) | Not Retained, N = 197<sup>1</sup> (49%) | Total, N = 418<sup>1</sup> | p-value<sup>2</sup> | N | Good Adherence, N = 141<sup>1</sup> (81%) | Poor Adherence, N = 34<sup>1</sup> (19%) | Total, N = 175<sup>1</sup> | p-value<sup>2</sup> |
| **Demographics** |  |  |  |  |  |  |  |  |  |  |  |  |  |  |  |
| Age Group | 623 |  |  |  |  | 418 |  |  |  |  | 175 |  |  |  |  |
| 25+ Years |  | 332 (58%) | 26 (49%) | 358 (57%) | 0.3 |  | 121 (55%) | 112 (57%) | 233 (56%) | 0.7 |  | 79 (56%) | 16 (47%) | 95 (54%) | 0.5 |
| Age in Years, Median (IQR) |  | 26 (22, 30) | 24 (22, 32) | 25 (22, 30) | 0.8 |  | 25 (22, 31) | 25 (23, 30) | 25 (22, 30) | 0.9 |  | 25 (22, 32) | 24 (22, 29) | 25 (22, 31) | >0.9 |
| Less than Grade 12 Education | 623 | 292 (51%) | 21 (40%) | 313 (50%) | 0.14 | 418 | 112 (51%) | 94 (48%) | 206 (49%) | 0.6 | 175 | 78 (55%) | 14 (41%) | 92 (53%) | 0.2 |
| Monthly income, Rand | 623 |  |  |  | 0.7 | 418 |  |  |  | 0.8 | 175 |  |  |  | 0.8 |
| No Income |  | 307 (54%) | 27 (51%) | 334 (54%) |  |  | 115 (52%) | 109 (55%) | 224 (54%) |  |  | 77 (55%) | 17 (50%) | 94 (54%) |  |
| R 5,000 and less |  | 224 (39%) | 21 (40%) | 245 (39%) |  |  | 90 (41%) | 75 (38%) | 165 (39%) |  |  | 54 (38%) | 14 (41%) | 68 (39%) |  |
| Over R 5,000 |  | 39 (6.8%) | 5 (9.4%) | 44 (7.1%) |  |  | 16 (7.2%) | 13 (6.6%) | 29 (6.9%) |  |  | 10 (7.1%) | 3 (8.8%) | 13 (7.4%) |  |
| Household size, including children, Median (IQR) | 621 | 4 (3, 5) | 3 (2, 6) | 4 (3, 5) | >0.9 | 416 | 4 (3, 6) | 4 (2, 5) | 4 (3, 5) | 0.004 | 174 | 4 (3, 5) | 5 (4, 6) | 4 (3, 6) | 0.066 |
| Number of living children, Median (IQR) | 623 | 1 (0, 2) | 0 (0, 1) | 1 (0, 2) | 0.11 | 418 | 1 (0, 1) | 1 (0, 2) | 1 (0, 2) | >0.9 | 175 | 1 (0, 2) | 1 (0, 1) | 1 (0, 2) | >0.9 |
| Currently married | 623 | 114 (20%) | 7 (13%) | 121 (19%) | 0.3 | 418 | 46 (21%) | 38 (19%) | 84 (20%) | 0.8 | 175 | 32 (23%) | 6 (18%) | 38 (22%) | 0.7 |
| Currently cohabiting (regardless of marital status) | 623 | 207 (36%) | 11 (21%) | 218 (35%) | 0.034 | 418 | 77 (35%) | 78 (40%) | 155 (37%) | 0.4 | 175 | 56 (40%) | 7 (21%) | 63 (36%) | 0.059 |
| **Pregnancy and Pregnancy Intentions** |  |  |  |  |  |  |  |  |  |  |  |  |  |  |  |
| Gestational Age in Weeks at Baseline | 620 |  |  |  |  | 429 |  |  |  |  | 175 |  |  |  |  |
| >= 20 Weeks |  | 318 (56%) | 31 (58%) | 349 (56%) | 0.8 |  | 115 (52%) | 124 (59%) | 239 (56%) | 0.2 |  | 70 (50%) | 24 (71%) | 94 (54%) | 0.045 |
| Gestational Age in Weeks, Median (IQR) |  | 21 (14, 29) | 22 (13, 31) | 21 (14, 29) | 0.8 |  | 20 (14, 27) | 23 (14, 32) | 21 (14, 29) | 0.055 |  | 19 (14, 27) | 24 (19, 30) | 20 (14, 27) | 0.045 |
|  | N | On PrEP, N = 570<sup>1</sup> (91%) | Not on PrEP, N = 53<sup>1</sup> (9%) | Total, N = 623<sup>1</sup> | p-value<sup>2</sup> | N | Retained, N = 221<sup>1</sup> (53%) | Not Retained, N = 197<sup>1</sup> (49%) | Total, N = 418<sup>1</sup> | p-value<sup>2</sup> | N | Good Adherence, N = 141<sup>1</sup> (81%) | Poor Adherence, N = 34<sup>1</sup> (19%) | Total, N = 175<sup>1</sup> | p-value<sup>2</sup> |
| Gravidity | 623 |  |  |  |  | 418 |  |  |  |  |  |  |  |  |  |
| Multigravida |  | 376 (66%) | 29 (55%) | 405 (65%) | 0.14 |  | 145 (66%) | 132 (67%) | 277 (66%) | 0.8 | 175 | 94 (67%) | 24 (71%) | 118 (67%) | 0.8 |
| Gravidity, Median (IQR) |  | 2 (1, 3) | 2 (1, 3) | 2 (1, 3) | 0.3 |  | 2 (1, 3) | 2 (1, 3) | 2 (1, 3) | 0.8 | 175 | 2 (1, 3) | 2 (1, 2) | 2 (1, 3) | 0.6 |
| Did not intent to get pregnant or changed intention | 622 | 396 (70%) | 32 (60%) | 428 (69%) | 0.2 | 418 | 149 (67%) | 134 (68%) | 283 (68%) | >0.9 | 175 | 94 (67%) | 26 (76%) | 120 (69%) | 0.4 |
| Did not want to have a baby or changed feelings | 623 | 378 (66%) | 29 (55%) | 407 (65%) | 0.12 | 418 | 144 (65%) | 127 (64%) | 271 (65%) | >0.9 | 175 | 89 (63%) | 26 (76%) | 115 (66%) | 0.2 |
| ***HIV Risk and Sexual Behaviors*** |  |  |  |  |  |  |  |  |  |  |  |  |  |  |  |
| Partner not tested for HIV or Don't Know Partner's HIV Status | 623 | 153 (27%) | 17 (32%) | 170 (27%) | 0.5 | 418 | 67 (30%) | 43 (22%) | 110 (26%) | 0.063 | 175 | 42 (30%) | 14 (41%) | 56 (32%) | 0.3 |
| Partner HIV Status | 623 |  |  |  | 0.7 | 418 |  |  |  | 0.007 | 175 |  |  |  | 0.4 |
| HIV negative |  | 409 (72%) | 36 (68%) | 445 (71%) |  |  | 149 (67%) | 154 (78%) | 303 (72%) |  |  | 95 (67%) | 19 (56%) | 114 (65%) |  |
| HIV positive |  | 8 (1.4%) | 0 (0%) | 8 (1.3%) |  |  | 5 (2.3%) | 0 (0%) | 5 (1.2%) |  |  | 4 (2.8%) | 1 (2.9%) | 5 (2.9%) |  |
| Don't Know |  | 153 (27%) | 17 (32%) | 170 (27%) |  |  | 67 (30%) | 43 (22%) | 110 (26%) |  |  | 42 (30%) | 14 (41%) | 56 (32%) |  |
| Has any STI (CT, NG, TV) | 619 | 202 (36%) | 14 (27%) | 216 (35%) | 0.3 | 417 | 81 (37%) | 71 (36%) | 152 (36%) | >0.9 | 175 | 43 (30%) | 22 (65%) | 65 (37%) | <0.001 |
| Chances of getting HIV in the next year | 622 |  |  |  | 0.11 | 418 |  |  |  | 0.9 | 175 |  |  |  | 0.2 |
| No chance at all |  | 360 (63%) | 26 (50%) | 386 (62%) |  |  | 150 (68%) | 128 (65%) | 278 (67%) |  |  | 98 (70%) | 18 (53%) | 116 (66%) |  |
| Small chance |  | 178 (31%) | 20 (38%) | 198 (32%) |  |  | 57 (26%) | 58 (29%) | 115 (28%) |  |  | 33 (23%) | 14 (41%) | 47 (27%) |  |
| Moderate chance |  | 24 (4.2%) | 5 (9.6%) | 29 (4.7%) |  |  | 10 (4.5%) | 8 (4.1%) | 18 (4.3%) |  |  | 7 (5.0%) | 2 (5.9%) | 9 (5.1%) |  |
| Great chance |  | 8 (1.4%) | 1 (1.9%) | 9 (1.4%) |  |  | 4 (1.8%) | 3 (1.5%) | 7 (1.7%) |  |  | 3 (2.1%) | 0 (0%) | 3 (1.7%) |  |
| At least one sexual partner during pregnancy | 622 | 556 (98%) | 49 (94%) | 605 (97%) | 0.2 | 418 | 216 (98%) | 195 (99%) | 411 (98%) | 0.5 | 175 | 139 (99%) | 33 (97%) | 172 (98%) | 0.5 |
| Sex frequency, last three | 607 |  |  |  | 0.7 | 411 |  |  |  | 0.7 | 172 |  |  |  | 0.2 |
|  | N | On PrEP,<br>N = 570<sup>1</sup><br>(91%) | Not on<br>PrEP, N<br>= 53<sup>1</sup><br>(9%) | Total, N =<br>623<sup>1</sup> | p-value<sup>2</sup> | N | Retained<br>, N =<br>221<sup>1</sup><br>(53%) | Not<br>Retained, N<br>= 197<sup>1</sup><br>(49%) | Total, N<br>= 418<sup>1</sup> | p-value<sup>2</sup> | N | Good<br>Adherenc<br>e, N =<br>141<sup>1</sup><br>(81%) | Poor<br>Adherence<br>, N = 34<sup>1</sup><br>(19%) | Total, N<br>= 175<sup>1</sup> | p-value<sup>2</sup> |
| months |  |  |  |  |  |  |  |  |  |  |  |  |  |  |  |
| Once a month or less often |  | 64 (12%) | 5 (9.8%) | 69 (11%) |  |  | 21 (9.7%) | 22 (11%) | 43 (10%) |  |  | 10 (7.2%) | 6 (18%) | 16 (9.3%) |  |
| 2-4 times a month |  | 277 (50%) | 25 (49%) | 302 (50%) |  |  | 114 (53%) | 100 (51%) | 214 (52%) |  |  | 74 (53%) | 18 (55%) | 92 (53%) |  |
| 5- 20 times per month |  | 171 (31%) | 19 (37%) | 190 (31%) |  |  | 67 (31%) | 55 (28%) | 122 (30%) |  |  | 45 (32%) | 7 (21%) | 52 (30%) |  |
| More than 20 times per month |  | 44 (7.9%) | 2 (3.9%) | 46 (7.6%) |  |  | 14 (6.5%) | 18 (9.2%) | 32 (7.8%) |  |  | 10 (7.2%) | 2 (6.1%) | 12 (7.0%) |  |
| **Stigma, Depression, Intimate Partner Violence (IPV) and Substance Use** |  |  |  |  |  |  |  |  |  |  |  |  |  |  |  |
| High Internalized Stigma | 623 | 35 (6.1%) | 28 (53%) | 63 (10%) | <0.001 | 418 | 7 (3.2%) | 13 (6.6%) | 20 (4.8%) | 0.2 | 175 | 4 (2.8%) | 1 (2.9%) | 5 (2.9%) | >0.9 |
| I feel ashamed of using PrEP |  | 14 (2.5%) | 7 (13%) | 21 (3.4%) | <0.001 |  | 4 (1.8%) | 5 (2.5%) | 9 (2.2%) | 0.3 |  | 2 (1.4%) | 0 (0%) | 2 (1.1%) | >0.9 |
| I feel embarrassed about using PrEP |  | 6 (1.1%) | 4 (7.5%) | 10 (1.6%) | 0.003 |  | 0 (0%) | 5 (2.5%) | 5 (1.2%) | 0.034 |  | 0 (0%) | 0 (0%) | 0 (0%) | >0.9 |
| I do not feel empowered to use PrEP |  | 3 (0.5%) | 16 (30%) | 19 (3.0%) | <0.001 |  | 1 (0.5%) | 1 (0.5%) | 2 (0.5%) | 0.065 |  | 0 (0%) | 1 (2.9%) | 1 (0.6%) | 0.4 |
| High Anticipated Stigma | 623 | 142 (25%) | 20 (38%) | 162 (26%) | 0.061 | 418 | 40 (18%) | 58 (29%) | 98 (23%) | 0.009 | 175 | 27 (19%) | 4 (12%) | 31 (18%) | 0.4 |
| I think people will give me a hard time if I tell them I am taking PrEP |  | 55 (9.6%) | 8 (15%) | 63 (10%) | 0.14 |  | 16 (7.2%) | 27 (14%) | 43 (10%) | 0.023 |  | 10 (7.1%) | 1 (2.9%) | 11 (6.3%) | 0.8 |
| I think people will judge me negatively if I take PrEP |  | 59 (10%) | 8 (15%) | 67 (11%) | 0.3 |  | 21 (9.5%) | 30 (15%) | 51 (12%) | 0.021 |  | 13 (9.2%) | 1 (2.9%) | 14 (8.0%) | 0.5 |
| I think I am at greater risk for physical violence or rape if I take PrEP |  | 20 (3.5%) | 3 (5.7%) | 23 (3.7%) | 0.027 |  | 9 (4.1%) | 8 (4.1%) | 17 (4.1%) | 0.7 |  | 6 (4.3%) | 1 (2.9%) | 7 (4.0%) | >0.9 |
| Depressed (EPDS Summary Score >=13) | 622 | 43 (7.5%) | 2 (3.8%) | 45 (7.2%) | 0.6 | 418 | 21 (9.5%) | 13 (6.6%) | 34 (8.1%) | 0.4 | 175 | 15 (11%) | 2 (5.9%) | 17 (9.7%) | 0.5 |
| Concern about sexual violence or rape in the next 3 months | 622 |  |  |  | 0.091 | 418 |  |  |  | 0.4 | 175 |  |  |  | 0.8 |
| Not concerned at all |  | 293 (51%) | 19 (37%) | 312 (50%) |  |  | 117 (53%) | 104 (53%) | 221 (53%) |  |  | 74 (52%) | 21 (62%) | 95 (54%) |  |
|  | N | On PrEP, N = 570<sup>1</sup> (91%) | Not on PrEP, N = 53<sup>1</sup> (9%) | Total, N = 623<sup>1</sup> | p-value<sup>2</sup> | N | Retained, N = 221<sup>1</sup> (53%) | Not Retained, N = 197<sup>1</sup> (49%) | Total, N = 418<sup>1</sup> | p-value<sup>2</sup> | N | Good Adherence, N = 141<sup>1</sup> (81%) | Poor Adherence, N = 34<sup>1</sup> (19%) | Total, N = 175<sup>1</sup> | p-value<sup>2</sup> |
| Somewhat concerned |  | 49 (8.6%) | 5 (9.6%) | 54 (8.7%) |  |  | 16 (7.2%) | 23 (12%) | 39 (9.3%) |  |  | 9 (6.4%) | 1 (2.9%) | 10 (5.7%) |  |
| Concerned |  | 43 (7.5%) | 8 (15%) | 51 (8.2%) |  |  | 14 (6.3%) | 14 (7.1%) | 28 (6.7%) |  |  | 9 (6.4%) | 1 (2.9%) | 10 (5.7%) |  |
| Very concerned |  | 185 (32%) | 20 (38%) | 205 (33%) |  |  | 74 (33%) | 56 (28%) | 130 (31%) |  |  | 49 (35%) | 11 (32%) | 60 (34%) |  |
| Experienced any Intimate Partner Violence | 622 | 72 (13%) | 3 (5.8%) | 75 (12%) | 0.2 | 418 | 23 (10%) | 19 (9.6%) | 42 (10%) | >0.9 | 175 | 16 (11%) | 4 (12%) | 20 (11%) | >0.9 |
| Emotional IPV |  | 61 (11%) | 2 (3.8%) | 63 (10%) | 0.2 |  | 19 (8.6%) | 16 (8.1%) | 35 (8.4%) | >0.9 |  | 14 (9.9%) | 3 (8.8%) | 17 (9.7%) | >0.9 |
| Physical IPV |  | 49 (8.6%) | 2 (3.8%) | 51 (8.2%) | 0.3 |  | 14 (6.3%) | 17 (8.6%) | 31 (7.4%) | 0.5 |  | 11 (7.8%) | 2 (5.9%) | 13 (7.4%) | >0.9 |
| Sexual IPV |  | 6 (1.1%) | 0 (0%) | 6 (1.0%) | >0.9 |  | 1 (0.5%) | 3 (1.5%) | 4 (1.0%) | 0.3 |  | 1 (0.7%) | 0 (0%) | 1 (0.6%) | >0.9 |
| Any Drug or Alcohol Use, 12 mo. | 622 | 274 (48%) | 18 (35%) | 292 (47%) | 0.086 | 418 | 113 (51%) | 90 (46%) | 203 (49%) | 0.3 | 175 | 77 (55%) | 14 (41%) | 91 (52%) | 0.2 |

<sup>1</sup>Statistics presented: n (col %); median (IQR)
<sup>2</sup>Statistical tests performed: chi-square test of independence; Wilcoxon rank-sum test; Fisher's exact test

### PrEP initiation at baseline

Ninety-one percent (91%, n=570) of n=623 pregnant women initiated PrEP at baseline. Cohabitation was significantly higher in the initiation sample among those on PrEP (36% vs. 21%; p=0.034). Those who did not initiate PrEP at baseline reported higher internalized stigma (53% vs. 6.1%; p<0.001) and higher anticipated stigma (38% vs. 25%; p=0.061). (Table I).

### PrEP retention at three months

Among the n=570 pregnant women who initiated PrEP at baseline, n=418 were eligible for a three-month follow up visit. By study analysis date, 53% (n=221) returned for a three-month follow up visit for PrEP retention. Household size was higher in the retention sample among those who returned (4, IQR: 3-6; p=0.004). Fewer participants who returned for the 3-month visit reported an HIV negative partner compared to HIV positive or unknown status (67% vs. 78%; p=0.007) in comparison to those who missed their visit or did not return. Baseline anticipated PrEP stigma was higher among those who missed or did not return for their 3-month visit, compared to those who returned (29% vs. 18%; p=0.009) (Table I).

### PrEP continuation at three months

Among the n=221 women who were retained at three months, 79.6% (n=176) continued on PrEP through the first quarterly follow up visit. There were no differences in demographics when comparing those who continued on PrEP with those who did not continue on PrEP in those who returned for the 3-month visit, so we did not tabulate these results.

### PrEP adherence at three months

Among the n=175 women who continued on PrEP through the first quarterly follow up visit, 81% (n=141) reported good adherence with a 30-day recall self-report measure. Poorly adherent participants in the adherence sample had significantly higher median gestational age study entry compared to those with good adherence (19 weeks vs 24 weeks; p=0.045) and had significantly higher baseline STI prevalence compared to those with good adherence (65% vs 30%; p<0.001). (Table I).

### Exploratory Factor Analysis: Internalized and Anticipated Stigma

We measured stigma with seven questions, of which six questions loaded into two separate factors (internalized PrEP stigma: α=0.811; anticipated PrEP stigma: α=0.777). Individual factor loadings are presented in Table II. Approximately 10% of participants in the initiation sample (n=63), 5% in the retention sample (n=21), 3% in the continuation sample (n=7) and 3% in the adherence sample (n=5) reported high internalized stigma. Prevalence of anticipated stigma was higher, with 26% (n=162) of participants in the initiation sample, 23% (n=98) in the retention sample, 18% (n= 40) in the continuation sample and 18% (n=31) the adherence sample reporting high anticipated stigma.

**Table II.** Factor loadings for two stigma scales.

| Survey question | Factor loading | $\alpha$ |
| --- | --- | --- |
| ***Factor 1: Internalized PrEP Stigma (sample units = 623)*** |  | **0.811** |
| I feel ashamed of using PrEP | 0.824 |  |
| I feel embarrassed about using PrEP | 0.859 |  |
| I do not feel empowered to use PrEP | 0.556 |  |
| ***Factor 2: Anticipated PrEP Stigma (sample units = 623)*** |  | **0.777** |
| I think people will give me a hard time (such as make fun of me, or talk badly about me) if I tell them I am taking PrEP | 0.979 |  |
| I think people will judge me negatively if I take PrEP | 0.721 |  |
| I think I am at greater risk for physical violence or rape if I take PrEP | 0.424 |  |
| ***Eliminated*** |  |  |
| People will think I am behaving responsibly by taking PrEP | -0.223 |  |

We hypothesized that internalized stigma and anticipated stigma are associated with lower PrEP initiation, retention, continuation and adherence and stand as barriers for pregnant PrEP users, and that partner HIV testing is an important factor in determining both anticipated stigma and PrEP initiation, retention, continuation and adherence. We tested this hypothesis with multivariable logistic regression with three outcomes: PrEP initiation at first antenatal visit, retention at 3 months after baseline, PrEP continuation at 3 months after baseline, and PrEP adherence at 3 months.

### Factors Independently Associated with PrEP Initiation, Continuation and Adherence

Reported internalized stigma was associated with lower odds of PrEP initiation at baseline (aOR: 0.06, 95% CI: 0.03-0.12) after controlling for gravidity, education and gestational age at baseline. Additionally, anticipated stigma was associated with lower odds of PrEP initiation at baseline (aOR: 0.53, 95% CI: 0.29-0.97) after controlling for gravidity, education, gestational age at baseline and partner HIV testing (Table III).

**Table III.**
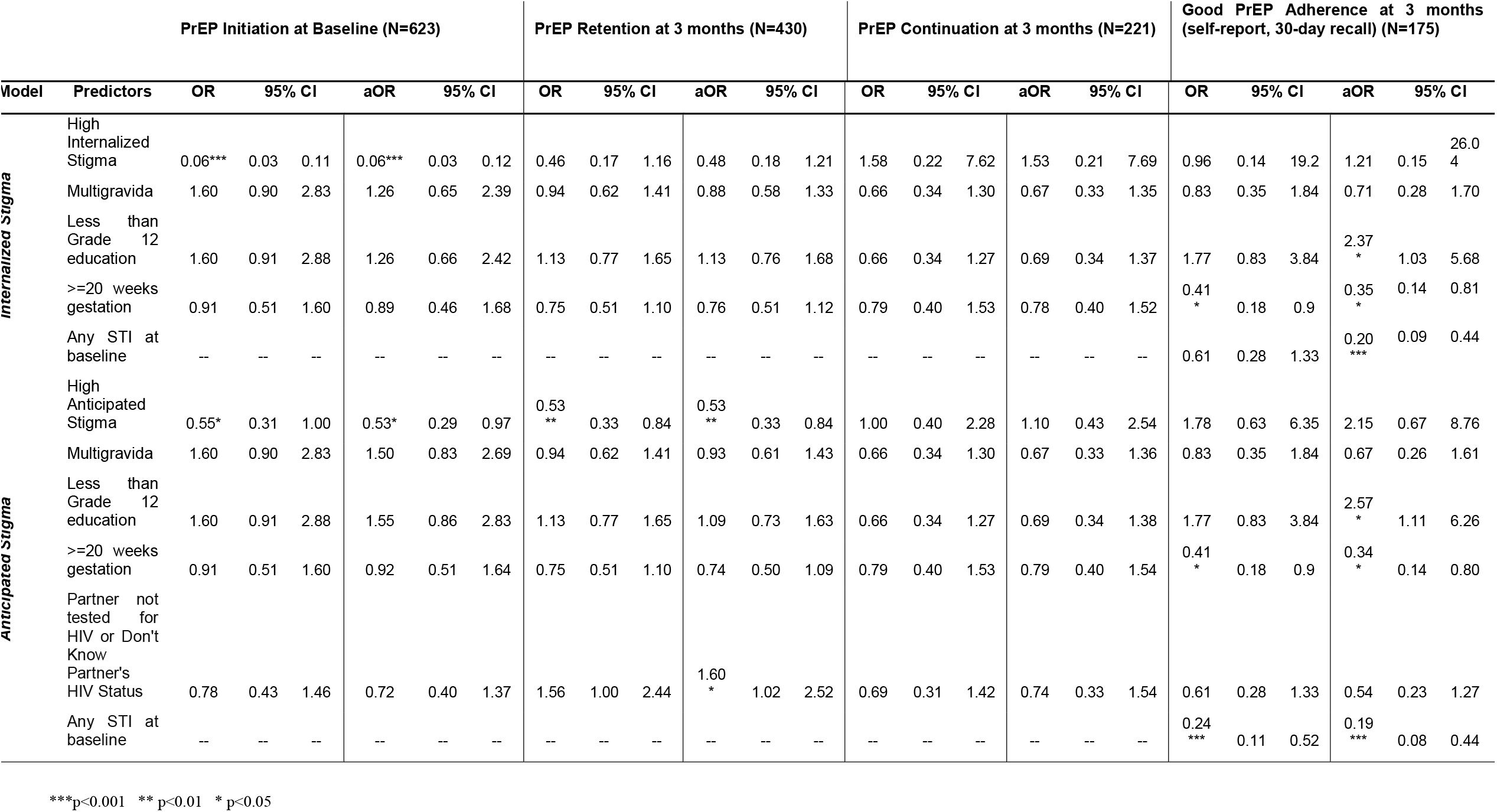
Crude and adjusted associations between outcomes of interest and selected stigma measures.

Anticipated stigma was associated with lower odds of returning for three-month study follow up and PrEP prescription among those who initiated PrEP at baseline (aOR: 0.53, 95% CI: 0.34-0.86) adjusting for covariates. Having a partner who had not tested for HIV or whose HIV serostatus was unknown were associated with higher odds of returning for the three-month follow up visit (aOR: 1.60, 95% CI: 1.02-2.52) after controlling for gravidity, education, gestational age at baseline (Table III). We did not find any association between those who continued on PrEP at three-month follow up compared to those who did not continue on PrEP.

Among those who continued on PrEP at three-month follow up, neither internalized stigma nor anticipated stigma were associated with higher adherence measured by self-report. Less than Grade 12 education was associated with higher adherence measured by self-report in the internalized stigma model (aOR: 2.37, 95% CI: 1.03-5.68) and the anticipated stigma model (aOR: 2.57, 95%CI: 1.11, 6.26). Additionally, we found that participants who entered the study with gestational age of at least 20 weeks were at lower odds of good adherence in both the internalized stigma model (aOR: 0.35, 95% CI: 0.14-0.81) and the anticipated stigma model (aOR: 0.34, 95% CI: 0.14-0.80). Finally, women entering the study with an STI had lower odds of good adherence compared to women without an STI in both the internalized stigma model (aOR: 0.20, 95% CI: 0.09-0.44) and the anticipated stigma model (aOR: 0.19, 95% CI: 0.08-0.44) (Table III).

## Discussion

We identified strong relationships between stigma and PrEP initiation at baseline and PrEP retention at three-months in a cohort of pregnant and postpartum women. We also identified an association between limited knowledge of partner serostatus and PrEP retention, and between both STI at baseline and gestational age at baseline and PrEP adherence, controlling for PrEP stigma. Specifically, we found that high PrEP internalized stigma was independently associated with lower odds of PrEP initiation, and that high anticipated PrEP stigma was independently associated both with lower odds of PrEP initiation at baseline and with lower odds of PrEP retention at three months. Additionally, PrEP users who did not know their partner’s serostatus prior to their first antenatal visit were at lower odds of returning for PrEP follow up at three months after controlling for anticipated PrEP stigma. Finally, women who had an STI at baseline or who entered the study with gestational age of at least 20 weeks were at lower odds of good PrEP adherence at three months after controlling for either internalized or anticipated PrEP stigma.

The results in this study build on previous assessments of stigma and PrEP initiation, retention, continuation and adherence in South Africa and elsewhere, in which stigma was a driver of PrEP discontinuation among women, MSM and sex workers. (7,18,21,37) While other efforts have categorized stigma as a barrier to PrEP initiation, retention, continuation and adherence, this study further describes internalized stigma and anticipated stigma as individual-level and relational-level barriers with distinct drivers and programmatic solutions. Prior studies demonstrated that stigma associated with the PrEP occasionally led to conflict with male partners as well as PrEP/study termination. Corneli et al. found that while participants reported that ART was potent and beneficial for HIV-infected individuals, they were regarded as potentially harmful when taken by HIV-negative individuals. (18)

Based on our results, internalized stigma is associated with lower PrEP initiation in pregnant women, while anticipated stigma is associated with lower PrEP initiation and PrEP retention. In contrast, HIV stigma measures generally showed no association between groups at initiation, retention or continuation. These findings highlight important differences in characterizing stigma among pregnant and post-partum women in South Africa. First, internalized PrEP stigma describes an individual’s perception of their own PrEP use as negative or shameful. (23) The relationship between high internalized stigma and lower PrEP initiation indicates that the barrier imposed by internalized stigma is highest at PrEP initiation and underscores the need for individual-level stigma reduction strategies. Second, anticipated stigma focuses on a person’s expectation of discrimination or prejudice because of their PrEP use. (23) The relationships between high anticipated stigma and both lower PrEP initiation and retention indicate that anticipated stigma serves as a continuous barrier for PrEP use which requires a different set of interventions outside of internalized stigma mitigation.

Additionally, there are other risk factors that were associated with sub-optimal PrEP use including presenting late for first antenatal care visit (>20 weeks), low educational attainment, and STI diagnosis at baseline. Reaching these high risk women will be essential to preventing HIV acquisition in pregnancy and postpartum periods and will require interventions such as flexible, differentiated care models that require less frequent clinic visits for prescriptions, HIV self-testing, and community or home delivery of PrEP. (17,38,39)

These findings are essential for improving PrEP use among pregnant and post-partum women in South Africa. Disclosure of PrEP use to partners and family members may also help to reduce levels of internalized and anticipated stigma. For example, having a partner who had not been tested for HIV or whose status was unknown was associated with lower odds of retention. This finding suggests that outside of anticipated stigma, other relational-level factors affect continued PrEP use over time and should be considered in the context of reducing anticipated stigma. Previous studies have identified that home-based HIV testing is a cost-effective intervention associated with higher uptake of couples testing, higher uptake of male partner testing and improved HIV status disclosure. (40–42) As high anticipated stigma is associated with lower PrEP initiation and retention, interventions which aim to engage male partners in HIV prevention through partner testing and counseling, awareness building and adherence support could reduce the consistent barrier faced by pregnant and post-partum women who experience high anticipated stigma.

This study is one of the first to describe stigma in a cohort of pregnant and postpartum women on PrEP in South Africa. There is little existing research on PrEP stigma among pregnant women and even fewer quantitative studies among these. Additionally, our validated scales for internalized PrEP stigma and anticipated PrEP stigma show high internal consistency and reliability for continued measurement and monitoring of stigma among pregnant and post-partum women in South Africa.

Despite these strengths, this study suffers from certain limitations; namely, its cross-sectional nature and self-reported adherence outcome. These cross-sectional associations indicate that PrEP retention at three months is lower among women with high anticipated stigma compared to those with low anticipated stigma, but future longitudinal analyses will be important to better understand these relationships over time. Additionally, adherence was self-reported and may be over reported due to social desirability bias or recall error. We will continue to assess the true levels of adherence from TFV-DP DBS in future analyses. Finally, the coronavirus disease 2019 (COVID-19) pandemic led to travel restrictions and government lockdown protocols which affected study retention due to site closures.(38,43,44)

## Conclusion

PrEP stands as an effective, safe strategy for primary HIV prevention among pregnant and post-partum women. Aside from programmatic and clinical barriers to optimize maternal PrEP use, behavioral factors act as important determinants of PrEP use and may act at both the individual and relational levels. Effective stigma mitigation must consider the level at which stigma originates (internalized, at the individual level or anticipated, at the relational and individual levels) to effectively reduce barriers for effective PrEP initiation and use. Improved awareness building among PrEP users and in the broader community can work to improve overall knowledge of PrEP and its benefits. Importantly, solutions which involve male partners – including HIV self-testing for partners, couples counseling and testing, and support in disclosing PrEP use by pregnant women– may be effective to reduce ongoing barriers to optimal PrEP use presented by anticipated PrEP stigma.

## Data Availability

Study data are stored securely per the approved protocol.

## Acknowledgements

We would like to thank our study participants, PrEP-PP study staff and the City of Cape Town Department of Health staff. DJD received funding from Fogarty International Center and National Institute of Health (K01TW011187), TC and LM received funding from National Institute of Mental Health (R01MH116771). The authors have no conflicts of interest to declare that are relevant to the content of this article.

