## Supplemental Material for "Maternal PrEP use in HIV-uninfected pregnant women in South Africa: Role of Stigma in PrEP initiation, retention and adherence"

### Directed Acyclic Graphs (DAGs)

We created two directed acyclic graphs (DAGs) to guide our analyses, presented in Figure S1. DAGs are labeled with PrEP Initiation (BL) as the outcome of interest for illustration, but also apply to the additional outcomes of PrEP retention at three months and PrEP continuation at three months. We propose that internalized stigma (Figure 2a) directly affects PrEP initiation at baseline, and is affected by gestational age at baseline, education level and gravidity. PrEP initiation at baseline is directly affected by gestational age at baseline, education level, gravidity, partner HIV testing and martial status. Based on the causal relationships defined in Figure S1a, we controlled for gestational age, education level and gravidity to minimize confounding from known sources and to measure the direct effect of internalized PrEP stigma on PrEP initiation at baseline. In Figure S1b, we propose that anticipated stigma directly affects PrEP initiation at baseline, and is affected by gestational age at baseline, education level, gravidity and partner HIV testing. PrEP initiation at baseline is directly affected by gestational age at baseline, education level, gravidity, partner HIV testing and marital status. Based on the causal relationships defined in Figure S1b, we controlled for gestational age, education level, gravidity and partner HIV testing to minimize confounding from known sources and to measure the direct effect of anticipated PrEP stigma on PrEP initiation at baseline. We propose that partner HIV status affects anticipated stigma (and not internalized stigma) because a partner’s unwillingness to be tested for HIV, or fear of partner’s unknown status, may affect a participant’s anticipation of stigma from her partner for taking her own steps to prevent HIV and to engage in HIV prevention, testing and care services. Aside from these causal differences, we did not examine internalized PrEP stigma and anticipated PrEP stigma together due to collinearity concerns (ρ = 0.5). For the PrEP adherence outcome, we also considered STI status at baseline under the premise that having an STI could affect PrEP stigma (internalized or anticipated) and could also affect ability or willingness to adhere to PrEP. In contrast, we decided that while STI at baseline could affect PrEP stigma (internalized or anticipated) it would not affect PrEP initiation, retention and continuation. These DAGs are shown in Figure S2. All DAGs were drawn using the R package ‘dagitty’. (45)

| a. | 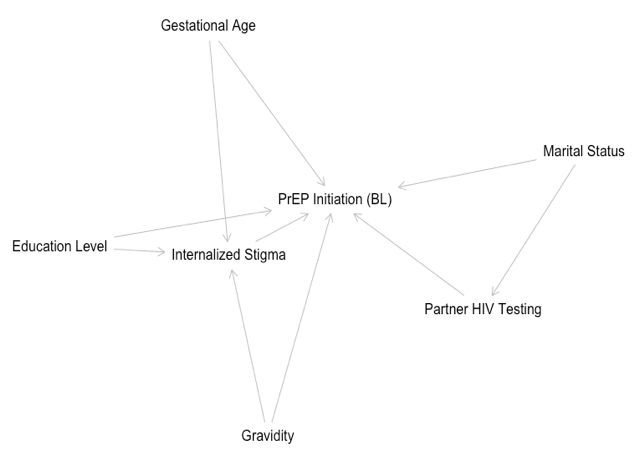 | b. | 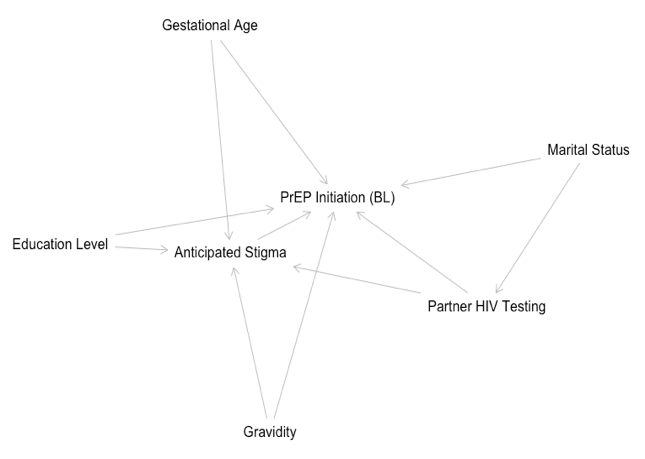 |
| --- | --- | --- | --- |
| **Figure S1.** Directed acyclic graphs (DAGs) for internalized PrEP stigma (a) and anticipated PrEP stigma (b) in the context of PrEP initiation, PrEP retention and PrEP continuation. | | | |

| a. | 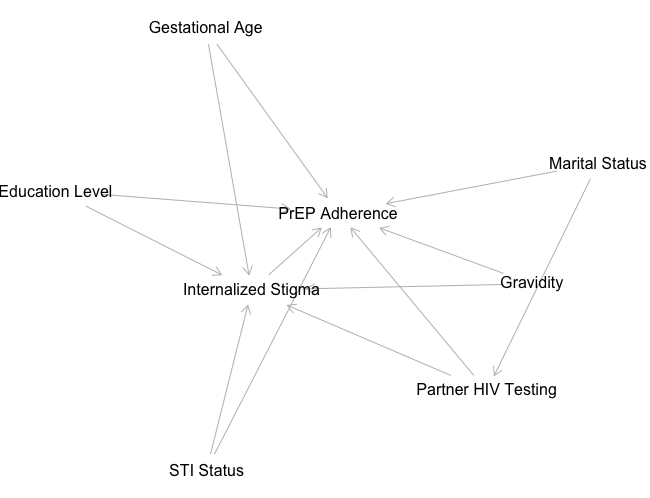 | b. | 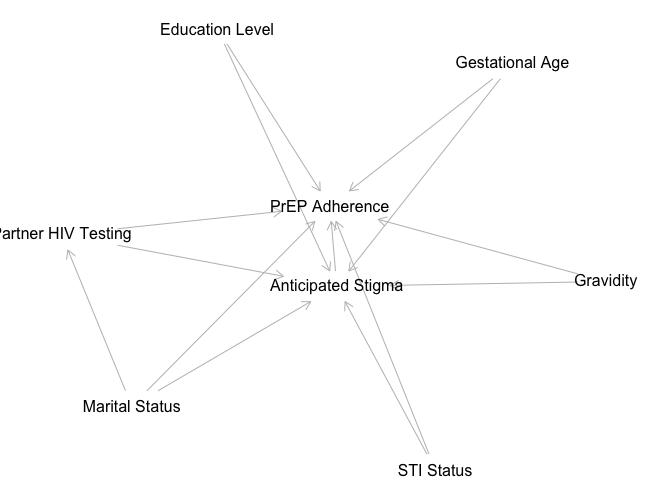 |
| --- | --- | --- | --- |
| **Figure S2.** Directed acyclic graphs (DAGs) for internalized PrEP stigma (a) and anticipated PrEP stigma (b) in the context of PrEP adherence. | | | |
